# Association Between Major Depressive Disorder and the Development of Cerebral Small Vessel Disease: A Mendelian Randomization Study

**DOI:** 10.1101/2023.11.21.23298814

**Authors:** Ting Lu, Lijun Luo, Jie Yang, Yueying Li, Daiyi Chen, Haiyang Sun, Huijuan Liao, Wen Zhao, Zhixuan Ren, Yan Xu, Shiyao Yu, Xiao Cheng, Jingbo Sun

**Author notes:** These three authors contributed equally to this work (Ting Lu, Lijun Luo, Jie Yang). **Corresponding authors:** Jingbo Sun (ORCID: 0000-0001-6609-5274) Department of Neurology, Guangdong Provincial Hospital of Traditional Chinese Medicine Guangzhou 510120, China Email address Xiao Cheng (ORCID: 0000-0002-8264-9713) Department of Neurology, Guangdong Provincial Hospital of Traditional Chinese Medicine Guangzhou 510120, China.

## Abstract

**Background:** Although observational studies indicate a complex, bidirectional association between major depressive disorder (MDD) and cerebral small vessel disease (CSVD), the results are frequently inconsistent. This study investigated the potential correlation of MDD with both CSVD clinical outcomes and imaging markers, utilizing a bidirectional Mendelian randomization (MR) study design.

**Methods:** Instrumental variables for both MDD and CSVD were extracted from the latest or most extensive genome-wide association study (GWAS) data available for each phenotype. Clinical outcomes and imaging markers of CSVD were defined using several parameters. The inverse variance weighting (IVW) method with additional sensitivity and heterogeneity analyses was used. Furthermore, a separate GWAS for depression was used to validate our significant findings.

**Results:** In the forward MR analyses, the genetically predicted risk of MDD was positively associated with two CSVD phenotypes showing microscopic white matter (WM) damage: mean diffusivity (IVW OR = 2.191, 95 % CI 1.226 to 3.917, p = 0.008) and WM-enlarged perivascular space (OR = 1.053 95 % CI 1.006 to 1.101, p = 0.026). The use of an independent database for depression yielded no significant risk of depression associated with these two CSVD traits. Furthermore, reverse MR analyses showed no evidence of reverse causality between MDD and an altered CSVD risk.

**Conclusions:** This study utilizing MR imaging findings supports a substantial causal association between MDD and CSVD-related indicators of impaired WM microstructure. It is necessary to exercise caution when extending these results to individuals with depression.

## Introduction

Major depressive disorder (MDD) is a global public health concern with a substantial socio-economic burden. Almost 20 % of individuals experience at least one episode of MDD during their lifetime (Kupfer, Frank, & Phillips, 2012). Cerebral small vessel disease (CSVD) comprises several syndromes affecting the small vessels of the brain. The most acute manifestations of CSVD are intracerebral hemorrhage (ICH) and small vessel ischemic stroke (SVS) (Markus & de Leeuw, 2023), and the principal neuroimaging markers include lacunar infarcts, white matter hyperintensities (WMH), brain microbleeds (BMBs), enlarged perivascular space (PVS), and cerebral atrophy (Chen et al., 2019). Interestingly, three decades of accumulated research evidence suggests a complex bidirectional relationship between MDD and CSVD, with WMH being the most extensively researched neuroimaging indicator.

WMH correlates with the occurrence of depressive symptoms (de Groot et al., 2000; Desmarais et al., 2021). Compared to the normal control group, patients with MDD exhibited greater severity of deep WMH signals (Brown, Lewine, Hudgins, & Risch, 1992). WMH severity is a crucial predictor of future depression risk in patients with CSVD (Park et al., 2015; Jaroonpipatkul et al., 2022; Ottavi, Pepper, Bateman, Fiorentino, & Brodtmann, 2023; F. Zhang, Ping, Jin, Hou, & Song, 2023). Specifically, those with extensive WMH at baseline are more likely to develop major depressive symptoms (Qiu et al., 2017). Increased mean diffusivity (MD) and decreased fractional anisotropy (FA) are two key indicators of assessing WM microstructural damage (Liao et al., 2013; Nobuhara et al., 2006; Taylor et al., 2004). Lower FA values in adolescents (Vulser et al., 2018) and higher MD values (Özel et al., 2023) are correlated with greater individual risk and occurrence of depressive symptoms, respectively.

Both lacunar infarcts (Direk et al., 2016; Özel et al., 2023) and BMBs (Direk et al., 2016; Xu et al., 2017) are associated with the occurrence of depressive symptoms and disorders. CSVD severity correlates with the diagnosis of depression before and after ICH (Castello et al., 2022). In patients with MDD, a significant correlation was observed between the number of traumatic events and overall PVS volume (Ranti et al., 2022). However, these observational studies may be subject to reverse causation and residual confounding factors, leading to conflicting results (Ahmed et al., 2022; Mewton et al., 2019). Therefore, the causative link between MDD and CSVD remains to be elucidated.

Mendelian randomization (MR) represents a potent, up-and-coming method that employs genetic variance as an instrumental variable (IV) to evaluate the causal impact of an exposure on a given outcome (Emdin, Khera, & Kathiresan, 2017; Sekula, Del Greco M, Pattaro, & Köttgen, 2016). Herein, we conducted a bidirectional two-sample MR analysis to comprehensively evaluate the relationship between MDD and clinical outcomes associated with CSVD (ICH or SVS, lobar ICH or SVS, and non-lobar ICH or SVS) and radiological markers of CSVD (WMH, FA, MD, BMBs, and PVS). In addition, we used a separate depression genome-wide association study (GWAS) to validate our significant findings.

## Materials and methods

### Study design

This study was conducted in accordance with the STROBE-MR checklist (Supplement file 1: Table S1) (Skrivankova et al., 2021). We described our rationale, study design, and procedures in Figure 1. All data used in this study were publicly available and therefore did not require ethics approval.

**Figure 1.**
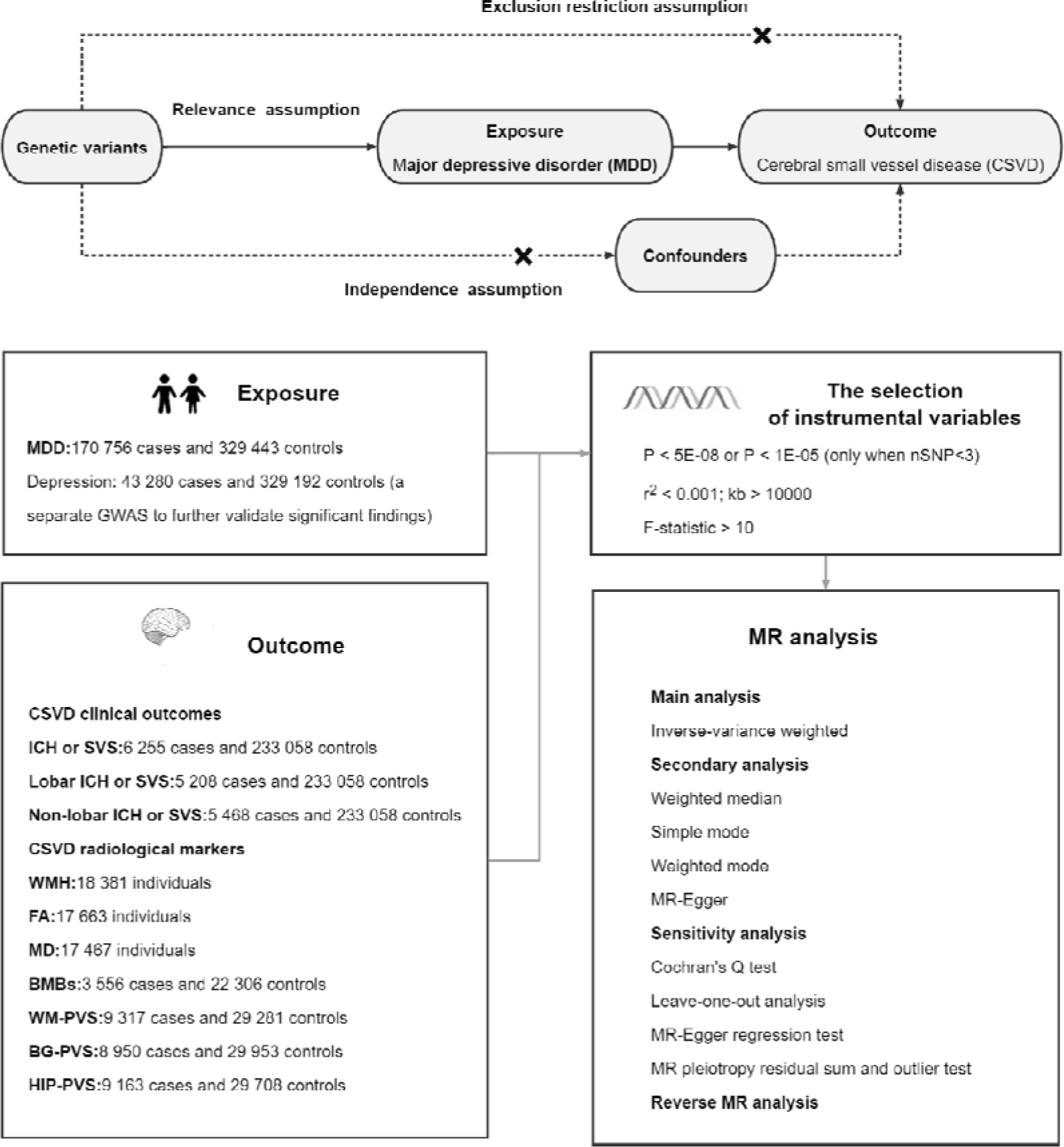
Diagrammatic overview of this MR study. The GVs selected as IVs met three core assumptions. (1) the relevance assumption: IVs should be strongly related to the exposure; (2) the independence assumption: the IVs should not be associated with other confounding factors; (3) the exclusion restriction assumption: the effect of the IVs on outcome should be exclusively through exposure. Abbreviations MDD, major depressive disorder; CSVD, cerebral small vessel disease; ICH, intracerebral hemorrhage; SVS, small-vessel ischemic stroke; WMH, white matter hyperintensities; FA, fractional anisotropy; MD, mean diffusivity; BMBs, brain microbleeds; WM, white matter; PVS, perivascular space; BG, basal ganglia; HP, hippocampal; nSNP, number of SNP; GVs, genetic variants; IVs, instrumental variables.

### Data sources

Details of the GWAS data sets are listed in Table 1. For MDD, we used the latest and largest GWAS summary statistics from the UK Biobank (UKB), which analyzed 170,756 MDD cases and 329,443 controls, and the Psychiatric Genomics Consortium (PGC) GWAS datasets, which analyzed 8,483,301 variants (Howard et al., 2019). The definition of MDD used by the UKB was based on self-reported symptoms, treatment, or electronic records (Howard et al., 2018). The definition of MDD used by the PGC was based on structured diagnostic interviews (Wray et al., 2018). To analyze depression, data were obtained from the FinnGen Study Release 9 of the GWAS for depression, F5_DEPRESSIO (Kurki et al., 2023), which included 43,280 patients and 329,192 controls. Depression was defined according to the International Classification of Diseases.

**Table 1.** Characteristics of the genome-wide association study used in this study.

|  | Ethnicity | Sample size | Adjusted | URL (data download) | Pubmed ID |
| --- | --- | --- | --- | --- | --- |
| **Exposure** |  |  |  |  |  |
| MDD | European | 170 756 cases and 329 443 controls | Sex, age, genotyping array, and the first 8 PCs | https://datashare.ed.ac.uk/handle/10283/3203 | 30718901 |
| Depression | European | 43 280 cases and 329 192 controls | Sex, age, genotyping batch, and 10 PCs | https://r9.risteys.finngen.fi/pheno/code/F5_DEPRESSIO | 36653562 |
| **Outcome** |  |  |  |  |  |
| ICH or SVS | European | 6 255 cases and 233 058 controls | Age, sex, and first 4 genetic PCs | https://cd.hugeamp.org/downloads.html | 31430377 |
| Lobar ICH or SVS | European | 5 208 cases and 233 058 controls | Age, sex, and first 4 genetic PCs | https://cd.hugeamp.org/downloads.html | 31430377 |
| Non-lobar ICH or SVS | European | 5 468 cases and 233 058 controls | Age, sex, and first 4 genetic PCs | https://cd.hugeamp.org/downloads.html | 31430377 |
| WMH | European | 18 381 individuals | Age at MRI, sex, genotyping array, the UK Biobank assessment center, the first 10 PCs, and MRI head motion indicators | https://cd.hugeamp.org/downloads.html | 32358547 |
| FA | European | 17 663 individuals | Age at MRI, sex, genotyping array, the UK Biobank assessment center, the first 10 PCs, and MRI head motion indicators | https://cd.hugeamp.org/downloads.html | 32358547 |
| MD | European | 17 467 individuals | Age at MRI, sex, genotyping array, the UK Biobank assessment center, the first 10 PCs, and MRI head motion indicators | https://cd.hugeamp.org/downloads.html | 32358547 |
| BMBs | Multiethnic (mainly European, 94%) | 3 556 cases and 22 306 controls | Age, sex, PCs, family, and relations (if applicable) | https://cd.hugeamp.org/downloads.html | 32913026 |
| WM-PVS | European | 9 317 cases and 29 281 controls | Age, sex, intracranial volume, PCs, and study site | https://www.ebi.ac.uk/gwas/studies/GCST90244151 | 37069360 |
| BG-PVS | European | 8 950 cases and 29 953 controls | Age, sex, intracranial volume, PCs, and study site | https://www.ebi.ac.uk/gwas/studies/GCST90244153 | 37069360 |
| HIP-PVS | European | 9 163 cases and 29 708 controls | Age, sex, intracranial volume, PCs, and study site | https://www.ebi.ac.uk/gwas/studies/GCST90244155 | 37069360 |

Abbreviations: GWAS, genome-wide association study; MDD, major depressive disorder; ICH, intracerebral hemorrhage; SVS, small-vessel ischemic stroke; WMH, white matter hyperintensities; FA, fractional anisotropy; MD, mean diffusivity; BMBs, brain microbleeds; WM, white matter; PVS, perivascular space; BG, basal ganglia; HP, hippocampal; PCs, principal components

The CSVD clinical outcomes were obtained from a meta-analysis of ICH GWAS results by location and SVS GWAS results from the MEGASTROKE study, which comprised “all location ICH or SVS,” “lobar ICH or SVS,” and “non-lobar ICH or SVS.” The study sample included 6,255 ICH or SVS cases and 233,058 controls of European ancestry (Chung et al., 2019).

The seven radiological markers of CSVD included WMH, FA, MD, BMBs, WM-PVS, basal ganglia (BG)-PVS, and hippocampal (HIP)-PVS.

WMH represents pathological changes in MRI scan results, whereas FA and MD demonstrate greater sensitivity to the disruption of normal function and structure than measuring pathology in isolation. GWAS data for three WM markers, WMH (N = 18,381), FA (N = 17,663), and MD (N = 17,467), were obtained from the UKB, Cohorts for Heart and Aging Research in Genomic Epidemiology (CHARGE), and WMH-Stroke multi-ethnic studies (Persyn et al., 2020). The WMH volume was measured using T1, T2, and fluid-attenuated inversion recovery sequences. FA and MD data were obtained from diffusion tensor imaging measurements of 48 brain regions.

The BMBs GWAS data were obtained from 11 population-based cohorts and 3 stroke cohort studies with case-control or case-only designs (Knol et al., 2020). BMBs were defined as small, low-signal lesions identifiable on a susceptibility-weighted imaging or T2-weighted gradient echo sequence. The study sample included 3,556 case and 22,306 control individuals, of whom 94 % were of European ethnicity.

The PVS data in European populations were obtained from a meta-analysis of 18 cohorts, including 9,317 case and 29,281 control individuals for WM-PVS, 8,950 case and 29,953 control individuals for BG-PVS, and 9,163 case and 29,708 control individuals for HIP-PVS (Duperron et al., 2023). PVSs were defined as fluid-filled spaces with the same signal as the cerebrospinal fluid and no high signal rim on T2-weighted or FLAIR sequences. The PVS burden was quantitatively defined by different scales, including the visual semiquantitative rating scale, among others.

### Selection of instruments

The filtering of instrumental variables (IVs) must fulfill three core assumptions (Figure 1). In the forward MR analysis, SNPs satisfying a genome-wide significance level of p < 5E-08 were initially selected (Zhang et al., 2023). For fewer than three remaining SNPs, a relaxed threshold of p < 1E-05 was used. Relaxing the statistical threshold for IVs has been employed in several MR studies in the psychiatric domain (Lee et al., 2023; Li et al., 2023; Pan et al., 2023). To exclude genetic variants with strong correlations, the clumping process was performed using the 1000 Genomes Project European reference panel (R2 < 0.001 and a window size of 10,000 kb). The SNPs and corresponding statistics were extracted from the CSVD GWAS dataset, deleting SNPs with minor allele frequency (MAF) < 0.01. Further, the MDD and CSVD data were harmonized by removing all palindromic SNPs. To satisfy the independence assumption, SNPs from the PhenoScannerV2 database that were significantly associated with confounding variables (Kamat et al., 2019) were collected. SNPs directly associated with CSVD were also excluded. We used the same rigorous criteria for selecting IVs in the reverse MR analysis. The variance explained (R^2^) by each SNP was calculated using the formula: R^2^ = 2 × MAF × (1−MAF) × beta^2^ (Si et al., 2021). The F-statistic was utilized to assess the strength of the genetic instrument. SNPs with F-statistic values < 10 (implying weak IVs) were removed from subsequent analyses (Burgess & Thompson, 2011).

### Statistical analysis

The primary analytic method used in this survey was the inverse variance weighting (IVW) method, which is known for its superior detection power (Burgess, Butterworth, & Thompson, 2013). Four other methods, namely weighted median, simple mode, weighted mode, and MR–Egger, were used for complementary analyses. For heterogeneity analysis, we used Cochran’s Q test (Greco M, Minelli, Sheehan, & Thompson, 2015) and leave-one-out analysis (Wu et al., 2021) to assess the level of heterogeneity among SNPs. The possible presence of pleiotropic effect was evaluated by the intercept of the MR–Egger regression test (Bowden, Davey Smith, & Burgess, 2015) and MR pleiotropy residual sum and outlier (MR-PRESSO) test (Verbanck, Chen, Neale, & Do, 2018). We used the same statistical analysis in the reverse MR analysis. We calculated odds ratios (ORs) to measure the causal effect between MDD and CSVD phenotypes. Considering the partial sample overlap between MDD and WMH in the UKB, we conducted a bias and type I error analysis to evaluate the potential bias risk attributable to sample overlap. Given the exploratory nature of this study, we did not perform correction of multiple comparisons.

The statistical software R (version 4.1.0, R Foundation for Statistical Computing, Vienna, Austria; https://www.R-project.org) was used to perform all statistical analyses and visualize the results obtained. The software packages were TwoSampleMR, MR-PRESSO, and Forestplot.

## Results

### Genetic instrument strength and sample overlap

For the 10 genetically related MDD-CSVD pairs, we only performed an MR analysis on pairs containing at least two qualifying IVs. In the forward MR analysis, all 10 pairs had SNPs available for analysis. In the reverse MR, two phenotypes, BG-PVS and HIP-PVS, had no IV remaining after the removal of the weak instrument (even when a loose threshold of p < 1E-05 was used); hence, they were not included in the MR analysis. The range of SNPs utilized as genetic instruments varied from 3 to 41, accounting for 0.16–23.63 % of the phenotypic variance, with F-statistics > 10 (range 20.92–1350.80) for all phenotypes, indicating the good strength of the IVs (Supplement file 1: Table S2). Details of all the SNPs included in the MR analysis are provided in the supplementary file (Supplement file 1: Table S3). The risk of bias due to sample overlap in the assessment was minimal (< 0.01, regardless of the overlap proportion).

### Causal relationship between MDD and CSVD

First, to comprehensively assess the effect of MDD on CSVD clinical outcomes and radiological markers, we used the IVW approach for the main analysis. Notably, as shown in the forest plot (Figure 2), we found a significant effect of MDD on two radiological markers of CSVD: MD and WM-PVS.

**Figure 2.**
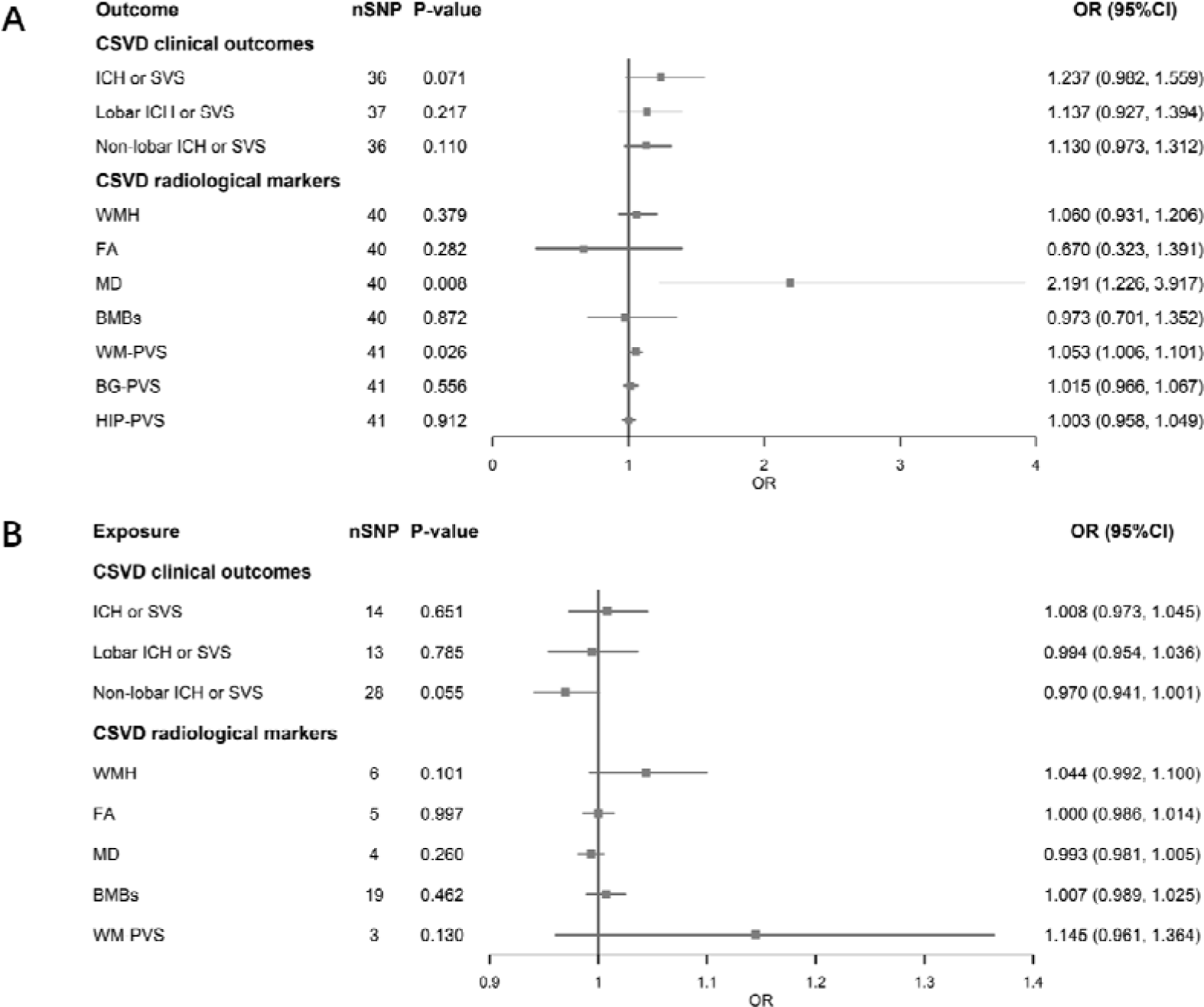
Bidirectional causal relationships between MDD and CSVD using the IVW approach. A The foward MR analyses examined the effect of MDD on CSVD clinical outcomes (n=3) and CSVD radiological markers (n=7). B The reverse MR analyses examined the effect of CSVD clinical outcomes (n=3) and CSVD radiological markers (n=4) on MDD. BG-PVS and HIP-PVS did not have a qualified IV available for analysis, even with a relaxed threshold (P=1E-05), and were therefore not included in the reverse MR analysis. Abbreviations nSNP, number of SNP; OR, odds ratio; CI, confidence interval; CSVD, cerebral small vessel disease; ICH, intracerebral hemorrhage; SVS, small-vessel ischemic stroke; WMH, white matter hyperintensities; FA, fractional anisotropy; MD, mean diffusivity; BMBs, brain microbleeds; WM, white matter; PVS, perivascular space; BG, basal ganglia; HIP, hippocampal.

The IVW approach demonstrated that a genetically determined higher risk of MDD was associated with increased MD (OR = 2.191, 95 % CI 1.226 to 3.917, p = 0.008). MR–Egger (OR = 97.532, 95 % CI 6.110 to 1556.906, p = 0.002) and weighted median (OR = 2.274, 95 % CI 1.117 to 4.631, p = 0.024) results were consistent with the IVW results (Figure 2). The MR–Egger intercept indicated the absence of pleiotropy (intercept = −0.116, p = 0.009) (Table 2), which may have caused the MR–Egger method to produce an OR with a wide range of results. Cochrane’s Q test showed no heterogeneity (Cochrane Q = 53.499, p = 0.061). The leave-one-out method showed no significant change in the IVW causal estimates after deleting any variant, suggesting the robustness of the results (Supplemental file 2: Figure S1). In addition, the MR-PRESSO test detected no significant outliers (p = 0.051).

**Table 2.** Results of the pleiotropy and heterogeneity test results for major depressive disorder (MDD) and cerebral small vessel disease (CSVD).

| Exposure | Outcome | nSNP | Cochrane's Q |  | MR Egger regression |  | MR PRESSO Global test |
| --- | --- | --- | --- | --- | --- | --- | --- |
|  |  |  | Q value | P value | intercept | P value | P value |
| MDD | ICH or SVS | 36 | 25.949 | 0.867 | 0.027 | 0.174 | 0.852 |
| MDD | Lobar ICH or SVS | 37 | 36.115 | 0.463 | 0.035 | 0.048 | 0.437 |
| MDD | Non-lobar ICH or SVS | 36 | 29.395 | 0.735 | 0.028 | 0.033 | 0.740 |
| MDD | WMH | 40 | 53.176 | 0.065 | -0.029 | 0.003 | 0.068 |
| MDD | FA | 40 | 88.348 | < 0.001 | 0.099 | 0.085 | 0.237 |
| MDD | MD | 40 | 53.499 | 0.061 | -0.116 | 0.009 | 0.051 |
| MDD | BMBs | 40 | 34.709 | 0.666 | 0.044 | 0.123 | 0.681 |
| MDD | WM-PVS | 41 | 47.365 | 0.197 | 0.001 | 0.844 | 0.210 |
| MDD | BG-PVS | 41 | 61.910 | 0.015 | 0.005 | 0.239 | 0.018 |
| MDD | HIP-PVS | 41 | 47.971 | 0.181 | -0.004 | 0.264 | 0.183 |
| ICH or SVS | MDD | 14 | 23.763 | 0.033 | 0.005 | 0.394 | 0.132 |
| Lobar ICH or SVS | MDD | 13 | 20.619 | 0.056 | -0.003 | 0.640 | 0.060 |
| Non-lobar ICH or SVS | MDD | 28 | 32.248 | 0.223 | -0.002 | 0.621 | 0.213 |
| WMH | MDD | 6 | 3.600 | 0.608 | -0.009 | 0.288 | 0.651 |
| FA | MDD | 5 | 4.693 | 0.320 | -0.009 | 0.281 | 0.408 |
| MD | MDD | 4 | 0.203 | 0.977 | -0.001 | 0.948 | 0.980 |
| BMBs | MDD | 19 | 30.089 | 0.037 | 0.003 | 0.562 | 0.046 |
| WM-PVS | MDD | 3 | 0.396 | 0.820 | -0.002 | 0.859 | NA |
| BG-PVS | MDD | 0 | NA | NA | NA | NA | NA |
| HIP-PVS | MDD | 0 | NA | NA | NA | NA | NA |

Abbreviations: nSNP, number of SNP; MDD, major depressive disorder; ICH, intracerebral hemorrhage; SVS, small-vessel ischemic stroke; WMH, white matter hyperintensities; FA, fractional anisotropy; MD, mean diffusivity; BMBs, brain microbleeds; WM, white matter; PVS, perivascular space; BG, basal ganglia; HIP, hippocampal; NA, not applicable.

A genetically determined higher risk of MDD was also associated with an increased risk of WM-PVS (OR = 1.053 95 % CI 1.006 to 1.101, p = 0.026) (Figure 2). The other four methods (MR–Egger, weighted median, simple mode, weighted mode) did not yield significant results (p > 0.05) (Supplemental file 1: Table S2). The sensitivity and heterogeneity analyses passed the test (all p > 0.05) (Table 2). The leave-one-out analyses indicated that the observed associations were stable (Supplemental 2: Figure S1).

For the other phenotypes, there was no evidence of an effect of MDD on the risk of clinical CSVD outcomes (all p > 0.05). There was also no evidence of an effect of MDD on the risk of WMH, FA, BMBs, BG-PVS, and HIP-PVS (Figure 2).

Depression data from the FinnGen database revealed no association between depression and the risk of MD (OR =1.084, 95 % CI 0.557 to 2.110, p = 0.812) and WM-PVS (OR = 0.872, 95 % CI 0.722 to 1.052, p = 0.151) by the IVW method (Supplemental 1: Table S4).

### Causal relationship between CSVD and MDD

In reverse MR, we examined the effect of eight phenotypes with qualified IVs on the risk of MDD. There was no evidence that ICH or SVS, lobar ICH or SVS, non-lobar ICH or SVS, WMH, FA, MD, BMBs, and WM-PVS affected the genetically predicted risk of MDD (Figure 2). Because there were only three SNPs in WM-PVS, the MR PRESSO Global test was not performed. No horizontal pleiotropy or heterogeneity (all p > 0.05) was found for the remaining phenotypes except for the BMBs (Cochrane Q = 30.089, p = 0.037, MR PRESSO Global test p = 0.046), suggesting that the results are reliable (Table 2).

## Discussion

To our knowledge, this is the first MR study to comprehensively investigate the causal association of MDD with CSVD clinical outcomes (ICH or SVS) and radiological markers (WMH, MD, FA, BMBs, and PVS). In forward MR, we found that the genetically predicted risk of MDD was positively associated with increased MD and the presence of WM-PVS. However, when the FinnGen dataset on depression was used, no genetically predicted risk of depression was found to be associated with these two CSVD traits (MD, WM-PVS), thus validating the study results. Additionally, reverse MR did not identify any causal association between CSVD risk and MDD. Since genetic variants are assigned to the offspring during meiosis, MR analyses can reflect lifelong exposure, reducing interference from reverse causation and confounding factors in comparison to observational studies. The aforementioned findings indicate a correlation between MDD and CSVD, implying that the genetically predicted risk of MDD could result in weakened microstructural integrity of cerebral WM. Consequently, curative measures or prevention of MDD may prove to be beneficial in averting future occurrences of CSVD.

Numerous epidemiological studies have investigated the intricate reciprocal association between MDD (or depression) and cerebral WM lesions (Gu et al., 2022; Pasi et al., 2016; Zhang et al., 2021). Patients with CSVD exhibiting depressive symptoms demonstrate a notable decrease in network efficiency and WM integrity compared to their counterparts without such symptoms (Lu et al., 2022; van Uden et al., 2015). The current MR study evaluated the mutually influential association of MDD with the WM macrostructure (WMH volume) and microstructure (FA and MD). Surprisingly, the analyses showed no evidence of a genetic association between MDD and WMH. This outcome is comparable to the results of two recent cohort studies: Clancy et al. discovered that heightened WMH was not linked to depression (2023), whereas Ali et al. illustrated that the load of WMH was not associated with depression 1 year after a stroke (2023). Another study, which implemented a machine learning algorithm, demonstrated that the severity of depression over time was unaffected by WMH volume (Ahmed et al., 2022).

Furthermore, the genetically predicted risk of MDD was associated with increased MD but not with FA. As WMH volume indicates the accumulation of WM damage over time, microstructural measurements may have greater sensitivity, implying that damage to microstructural integrity precedes WMH. The disparity between the two microscopic indicators could be attributed to MD being linked to blood-brain barrier (BBB) leakage, which occurs earlier in WMH progression and may be more receptive to identifying minor injuries. Conversely, FA seems to be affected more directly by myelin changes and may identify more severe injuries (Cubon, Putukian, Boyer, & Dettwiler, 2011; O’Dwyer et al., 2011; Song et al., 2003).

This study demonstrated that the MDD correlated positively with another form of WM microstructure damage: the WM-PVS. A possible explanation relates to perturbations in the central nervous system microenvironment in patients with MDD, including impaired BBB permeability (Yarlagadda, Alfson, & Clayton, 2009), leading to the recruitment and accumulation of blood immune cells and immune mediators around blood vessels (Winkler et al., 2021), resulting in PVS enlargement (Wuerfel et al., 2008).

A prior MR study explored the causal link between MDD and ischemic stroke, specifically SVS, and found that MDD was associated with a higher risk of SVS (Cai et al., 2019). We used a cross-phenotypic GWAS (Chung et al., 2019) and found no effect of genetically predicted risk of MDD on CSVD clinical phenotypes (ICH or SVS, lobar ICH or SVS, non-lobar ICH or SVS). One possible explanation for this discrepancy in results is that Cai et al.’s study used a looser genome-wide significance threshold (p < 1E-06) to allow the inclusion of more SNPs that contributed to MDD risk. In contrast, we used a more stringent threshold of p < 5E-08 when examining the three pairwise associations between MDD and CSVD clinical phenotypes. Furthermore, a recent MR in 2023 examined the risk association between MDD and ICH and found no causal relationship (Wang et al., 2022). Because we used larger and more comprehensive GWAS datasets for both MDD and CSVD clinical phenotypes, our findings may provide more reliable evidence of an association between the two.

Our study had some limitations. First, the positive results regarding the causal relationship between MDD and CSVD were not replicated with the FinnGen depression data. This suggests an association between MDD rather than depression and increased susceptibility to CSVD; therefore caution is needed in extending the results to individuals with depression who do not meet clinical diagnostic criteria for MDD.

Second, the association between CSVD and late-life depression (LLD), based on the “vascular depression hypothesis,” has been extensively studied previously (Aizenstein et al., 2016; Alexopoulos et al., 1997). However, in the current study, the lack of high-quality GWAS data on LLD limited more precise analyses of the association between depressive subtypes and CSVD. Future studies are expected to explore the complex bidirectional relationship between CSVD and depression subtypes using more extensive GWAS data.

Third, women have a significantly higher prevalence of depression (Kessler, McGonagle, Swartz, Blazer, & Nelson, 1993; Kuehner, 2017). An observational study found a significant association of depressive symptoms with periventricular WMH, deep WMH, and the number of asymptomatic infarcts in women but not in men (Wendell et al., 2010). Although the original GWAS dataset used was adjusted for sex, sex-stratified data were lacking. Further future studies that stratify differential effects by sex would be valuable.

Finally, despite the various sensitivity analyses, the possibility of uncontrolled pleiotropy or heterogeneity cannot be excluded. More large-scale prospective studies are needed in the future to verify the causal relationship between MDD and CSVD.

## Conclusion

In conclusion, the association between the risk of MDD and CSVD may have far-reaching physiological value and clinical implications, as identified in this MR study. The implication of this potential association involves the proactive identification of individuals at risk for the development and progression of CSVD, leading to the implementation of prevention strategies and improved clinical outcomes.

## Data Availability

All data generated or analyzed in this study are available in this published article and its supplementary information files.

## Ethical standards

Not applicable.

## Author contributions

TL, XC, and JBS conceptualized the study design. TL, LJL, JY, YYL, DYC, HYS, and HJL analyzed the data. WZ, ZXR, YX, and SYY verified the accuracy of the data. TL, LJL, and JY were major contributors to the writing of the manuscript. All authors read and approved the final version of the manuscript.

