## Supplement file 2 for "Association Between Major Depressive Disorder and the Development of Cerebral Small Vessel Disease: A Mendelian Randomization Study"

**Figure S1.** Causal relationship between MDD and two radiological markers of CSVD (MD and WM-PVS). **A** Scatter plot of the causal relationship between MDD and MD using five MR methods. **B** Leave-one-out plot of the causal relationship between MDD and MD. No significant change in the IVW causal estimates after deletion of any variant, suggesting that the results are robust. **C** Scatter plot of the causal relationship between MDD and WM-PVS. **D** Leave-one-out plot of the causal relationship between MDD and WM-PVS. **Abbreviations** MDD, major depressive disorder; CSVD, cerebral small vessel disease; MD, mean diffusivity; WM, white matter; PVS, perivascular space.

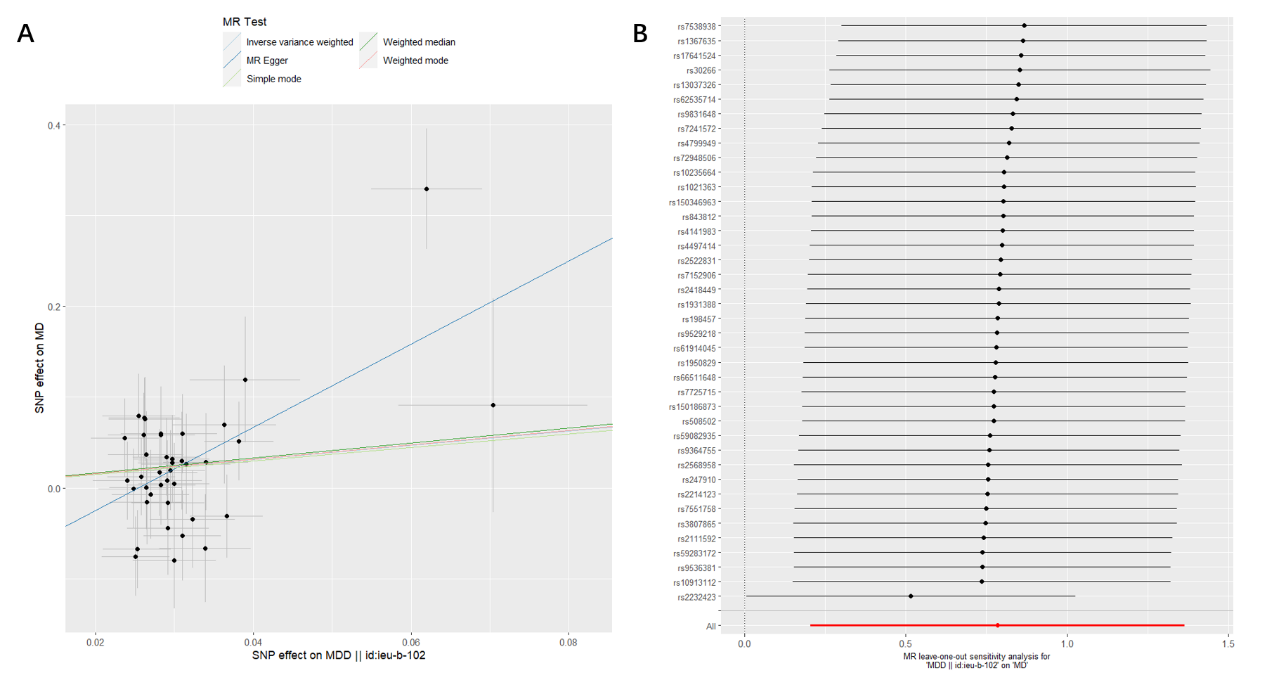

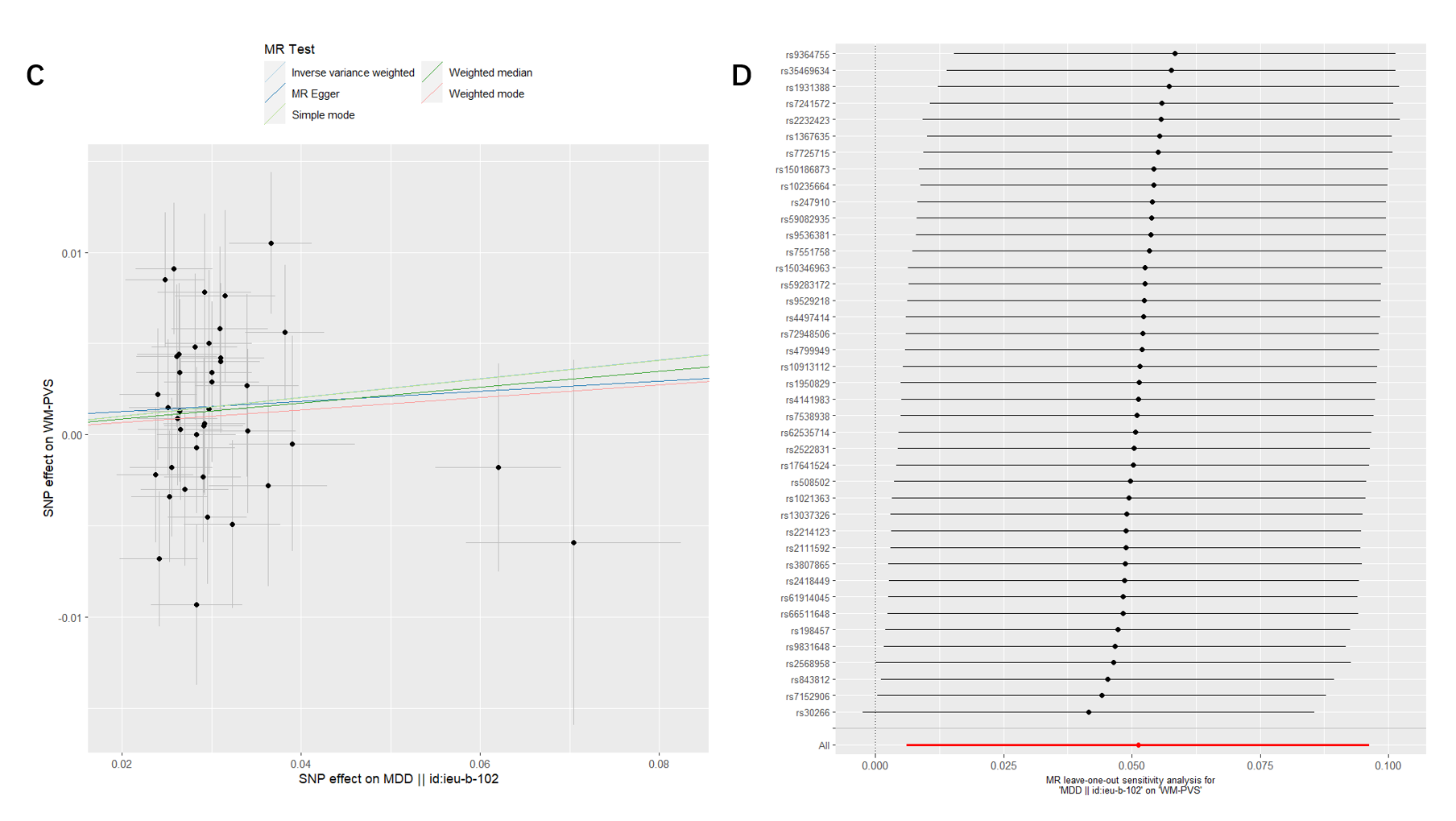
